# Severe acute infection and chronic pulmonary disease are risk factors for developing post-COVID-19 conditions

**DOI:** 10.1101/2022.11.30.22282831

**Authors:** Pritha Ghosh, Michiel J.M. Niesen, Colin Pawlowski, Hari Bandi, Unice Yoo, Patrick J. Lenehan, Praveen Kumar M., Mihika Nadig, Jason Ross, Sankar Ardhanari, John C. O’Horo, AJ Venkatakrishnan, Clifford J. Rosen, Amalio Telenti, Ryan T. Hurt, Venky Soundararajan

**Author notes:** Correspondence to: Ryan Hurt and Venky Soundararajan. These authors contributed equally (Pritha Ghosh, Michiel J.M. Niesen).

## Abstract

Post-COVID-19 conditions, also known as “long COVID”, has significantly impacted the lives of many individuals, but the risk factors for this condition are poorly understood. In this study, we performed a retrospective EHR analysis of 89,843 individuals at a multi-state health system in the United States with PCR-confirmed COVID-19, including 1,086 patients diagnosed with long COVID and 1,086 matched controls not diagnosed with long COVID. For these two cohorts, we evaluated a wide range of clinical covariates, including laboratory tests, medication orders, phenotypes recorded in the clinical notes, and outcomes. We found that chronic pulmonary disease (CPD) was significantly more common as a pre-existing condition for the long COVID cohort than the control cohort (odds ratio: 1.9, 95% CI: [1.5, 2.6]). Additionally, long-COVID patients were more likely to have a history of migraine (odds ratio: 2.2, 95% CI: [1.6, 3.1]) and fibromyalgia (odds ratio: 2.3, 95% CI: [1.3, 3.8]). During the acute infection phase, the following lab measurements were abnormal in the long COVID cohort: high triglycerides (mean_longCOVID_: 278.5 mg/dL vs. mean_control_: 141.4 mg/dL), low HDL cholesterol levels (mean_longCOVID_: 38.4 mg/dL vs. mean_control_: 52.5 mg/dL), and high neutrophil-lymphocyte ratio (mean_longCOVID_: 10.7 vs. mean_control_: 7.2). The hospitalization rate during the acute infection phase was also higher in the long COVID cohort compared to the control cohort (rate_longCOVID_: 5% vs. rate_control_: 1%). Overall, this study suggests that the severity of acute infection and a history of CPD, migraine, CFS, or fibromyalgia may be risk factors for long COVID symptoms. Our findings motivate clinical studies to evaluate whether suppressing acute disease severity proactively, especially in patients at high risk, can reduce incidence of long COVID.

## Introduction

According to CDC estimates, approximately 58% of the United States population has had a SARS-CoV-2 infection at least once through February 2022,^1^ and the total number of confirmed COVID-19 deaths surpassed 1 million in May 2022.^2^ Given the high prevalence of COVID-19 and its large burden on health systems and society overall, it is a public health imperative to understand the short, medium, and long-term effects of this disease so that optimal care can be offered to COVID-19 patients during their infection and their convalescence. There is mounting evidence that SARS-CoV-2 infection may have significant long-term health effects for some individuals. For example, some individuals, particularly those infected with earlier variants of SARS-CoV-2, may experience persistent loss of taste and/or smell.^3^ The WHO developed a clinical case definition for post-COVID-19 conditions (also known as “long COVID”), which include fatigue, shortness of breath, and cognitive dysfunction as common symptoms.^4^ In October 2021, an ICD code for long COVID was adopted internationally (U09.9). According to the National Center for Health Statistics (NCHS) Household Pulse Survey, approximately 34% of individuals who were infected with COVID-19 report symptoms lasting three months or more after their infection.^5^ One large retrospective study found that anosmia, hair loss, sneezing, ejaculation difficulty, and reduced libido were the most commonly reported long COVID symptoms, and risk factors include female sex, belonging to an ethnic minority, socioeconomic deprivation, smoking, obesity, and a wide range of comorbidities.^6^ Currently, prospective studies are underway to characterize the long-term sequelae of COVID-19, including the CDC INSPIRE study,^7^ and the NIH RECOVER initiative.^8^

Here, we conduct a large-scale retrospective analysis of de-identified electronic health records from a multi-state health system to characterize long COVID conditions and associated risk factors. We consider a cohort of patients with long COVID based on an ICD code diagnosis and a control cohort of COVID-19 patients without long COVID diagnosis. We perform 1:1 matching to ensure that the cohorts are balanced on clinical characteristics, including demographics, date of infection, geography, and the number of prior laboratory testing encounters. We examined trends in lab test measurements for these two matched cohorts during a baseline phase before COVID-19 diagnosis and an acute COVID-19 phase. In addition, we compared other clinical features between these two cohorts including hospitalization, diagnoses, medications, and signs and symptoms captured in clinical notes.

## Methods

### nference platform with de-identified electronic health record data

We used the nference Clinical nSights platform to conduct this analysis. This platform includes de-identified records from over 6.9 million patients, spanning multiple US states. This de-identified environment includes structured tables derived from electronic health records (EHR) data such as ECG waveforms, diagnosis codes, laboratory tests, vital signs, medications administered, medications ordered, procedures, and flowsheets. In addition, this environment includes unstructured tables derived from the EHR, such as ECG, radiology and pathology reports, and clinical notes. All personally identifiable information in this environment (e.g., names, locations, dates) have been excluded or substituted using a best-in-class de-identification methodology.^9^

### Study design

In the de-identified EHR database, the study population included all individuals with at least one positive SARS-CoV-2 PCR test between June 1, 2021 (four months before the first use of the long COVID ICD-10 code) to May 28, 2022. Individuals without a primary care provider on record in the health system or with no clinical encounters recorded in the past three years were excluded from the analysis. Individuals with at least one ICD-10 code for long COVID (U09.9, “Post COVID-19 condition, unspecified”) at least 7 days after a positive SARS-CoV-2 PCR test were grouped into the “Long COVID” cohort, and the rest of the study population without this ICD-10 code was grouped into the control cohort. For individuals in the long COVID cohort, the date of the most recent positive PCR test prior to the first U09.9 ICD-10 code was considered to be the index date. For individuals in the control cohort, the date of the first positive PCR test during the study period was considered to be the index date. In **Figure 1**, we provide an overview of the study design.

**Figure 1:**
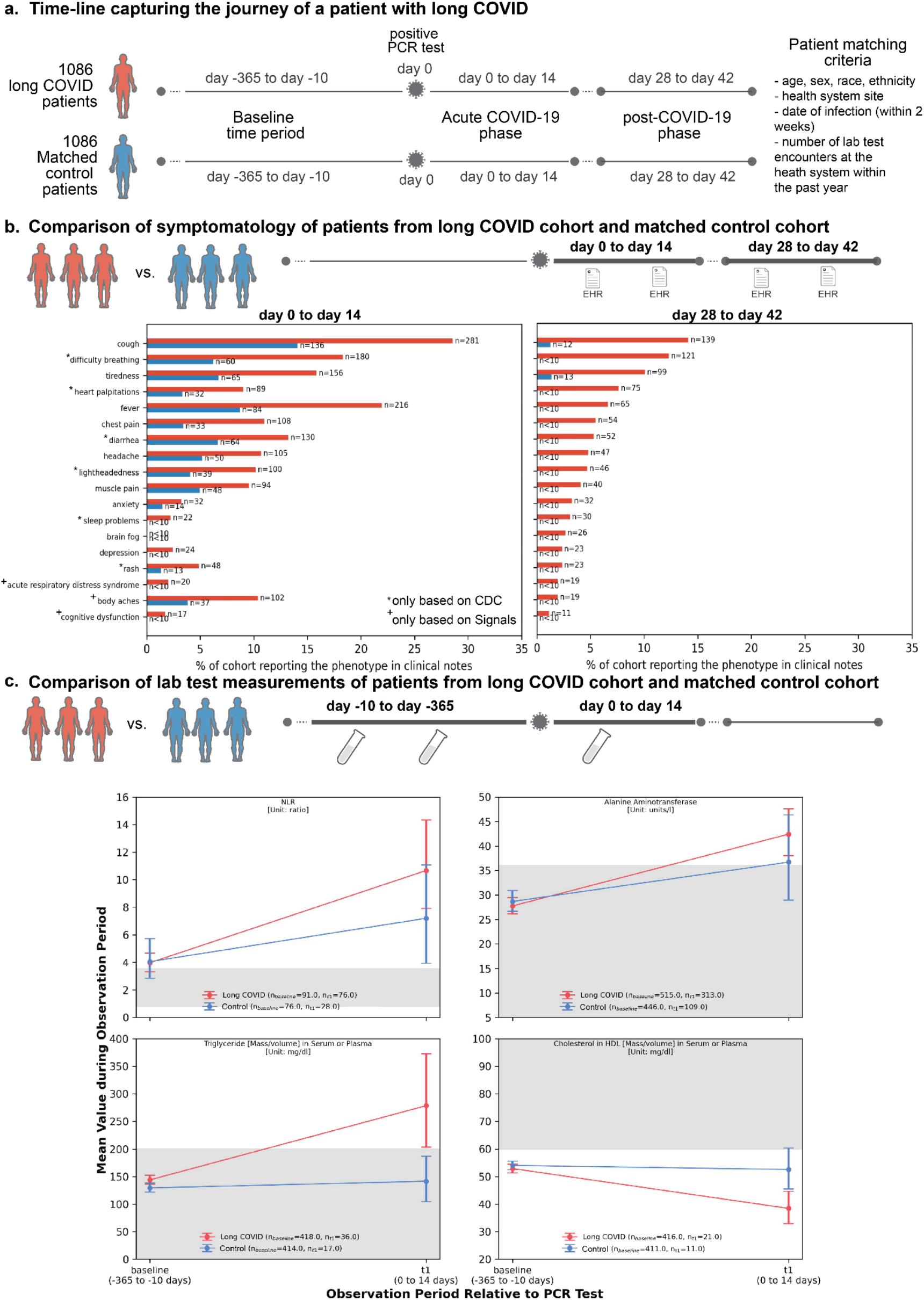
Study Overview. **(a)** Timeline capturing the journey of a patient with long COVID. There are three main phases — i) baseline (10 to 365 days before infection), ii) acute COVID-19 (0 to 14 days after infection), and iii) post-COVID-19 (28 to 42 days after infection). **(b)** Comparison of new onset symptoms and diseases recorded in EHR notes following a positive SARS-CoV-2 PCR test. Only phenotypes for which there is a significant difference in reporting (Fisher’s exact test, p-value < 0.05) between the long COVID and control cohorts are shown. **(c)** Lab test with significant difference between the matched long COVID vs. control cohorts. For each lab test, mean test values and 95% confidence intervals are shown. The normal ranges for these lab tests^28–31^ are shaded in gray.

### Definition of the matched control cohort

To identify risk factors for long COVID, we constructed a 1:1 matched control cohort starting from the unmatched study population. This cohort was exactly matched on potentially confounding factors for long COVID ICD-10 diagnosis, including demographics (age, sex, race, ethnicity), health system site, date of infection (within two weeks), and the number of lab test encounters at the health system within the past year. Individuals in the long COVID cohort without a corresponding matched control (54 out of 1,140 individuals) were dropped from the matched analysis.

### Extraction of phenotypes from clinical notes

A Bidirectional Encoder Representations from Transformers (BERT)-based classification model was used to classify the sentiment for phenotypes mentioned in EHR clinical notes. BERT is a transformer-based machine learning model used for natural language processing of unlabeled data. This model was previously used to identify signs and symptoms of COVID-19,^10^ short and long-term complications of COVID-19,^11^ and adverse events of mRNA-based COVID-19 vaccines.^12^ Given a sentence that includes any phenotype, this model outputs one of the following labels: “Yes” - confirmed diagnosis, “Maybe” - possible diagnosis, “No” - ruled out the diagnosis, or “Other” - none of the above. A dataset of 18,490 manually annotated sentences extracted from EHR clinical notes containing over 250 different phenotypes was used to train the model. The classification model achieves an out-of-sample accuracy of 93.6% and precision and recall values above 95%.^10^

For this study, we applied the BERT model to classify the sentiment of 64 phenotypes (**Table S1**) in the clinical notes for individuals in the long COVID and control cohorts during each of the study phases. This list of phenotypes was obtained from the CDC website for long COVID^13^ and publicly available literature sources, and the methodology to identify candidate long COVID phenotypes from publicly available literature sources is described in the following methods section. For the analysis of clinical notes, we first define the following phases: the baseline or pre-COVID-19 phase (10 to 365 days before infection), the acute COVID-19 phase (0 to 14 days after infection), and the post-COVID-19 phase (28 to 42 days after infection). The time window for the post-COVID-19 phase was selected to both capture most new long COVID diagnosis (60% of the long COVID cohort was diagnosed with long COVID before day 42, and 18% was diagnosed between day 28 to 42) and because during this time window we observed significant differences in overall phenotype reporting (**Figure S2**). Individuals without at least one clinical note during the baseline phase and individuals with less than 42 days of follow-up post-PCR were excluded from this analysis. For the baseline phase, an individual was counted as positive for the phenotype if they had at least one mention of the phenotype with a “Yes” label and the confidence score was greater than 0.8 (a “positive sentiment”). For each prediction, the confidence score is a number between 0 and 1 which reflects the certainty of the model that the prediction is correct, with 0 being the least certain and 1 being the most certain. In this study, we selected a threshold of 0.8 for the confidence score based on manual review of a subset of model predictions. For the acute and post-COVID-19 phases, an individual was counted as positive for a phenotype only if they had a positive sentiment for the phenotype during that phase (i.e. “Yes” label and confidence score > 0.8) without any positive sentiment in the baseline phase. We term such phenotypes as “new onset”. We have also quantified the overall prevalence of positive sentiments for any of the 64 phenotypes during 7-day intervals from 42 days before the positive PCR test to 42 days after the positive PCR test (**Figure S2**).

### Identification of candidate long COVID phenotypes from publicly available literature sources

The nferX Signals application (https://research.nferx.com/dv/202011/signals/) was used to determine candidate long COVID phenotypes from publicly available literature sources. This application enables the user to search for biomedical associations in free-text over 100 million documents from over 80K sources including but not limited to: PubMed articles, clinicaltrials.gov, patent applications, SEC filings, blogs, conferences, and news articles. For this study, we used this application to identify disease phenotypes frequently mentioned in the biomedical literature in the context of long COVID and its associated synonyms (e.g. “pasc”, “post-COVID condition”). The full list of disease phenotypes that were considered includes approximately 140K unique phenotypes compiled from 9 sources which are available in the nferX “Diseases” collection (see **Table S2**). The synonym lists for each of these phenotypes and “long COVID” were determined by the nferX Signals application. For each phenotype in the “Diseases” collection, we computed a metric called the “nferX local score” which measures the strength of the association between that phenotype and long COVID in the nferX corpus of biomedical literature. The formula to compute the nferX Local Score is provided in the Supplemental Materials (see **Figure S1**). In particular, phenotypes which co-occur relatively frequently with long COVID in the corpus within a specified word span achieve high local scores, and phenotypes which co-occur relatively infrequently achieve low local scores. Phenotypes with a significantly high association (local score > 3.0) for a word span of +/- 50 words were considered as candidate long COVID phenotypes, excluding COVID-19 and non-specific disease phenotypes. The final list of 64 phenotypes considered for this study is the union of the list of phenotypes on the CDC website for long COVID^13^ and the list of candidate long COVID phenotypes identified by the nferX Signals application (see **Table S1**).

### Comparison of lab measurements

For the matched long COVID and control cohorts, we computed: (a) the mean values of a lab test for each patient contributing to the analysis of this lab test (mean_individual_: patient-level data summarization), and (b) the mean values of a lab test for all mean_individual_ values of patients in a cohort (mean_population_: population-level data summarization). We performed these calculations for the baseline and acute COVID-19 phases. We compared the mean_population_ (hereafter referred to as ‘mean’) values for a lab test between — i) the baseline and acute COVID-19 phases for the long COVID cohort, and ii) the long COVID and control cohorts in the acute COVID-19 phase. In both cases, we report p-values from Mann-Whitney U tests, subsequently corrected for multiple comparisons using the Benjamini-Yekutieli (False Discovery Rate) method. We also calculated 95% confidence intervals around the mean_population_ values by bootstrap resampling (1000 samples).

### Comparison of clinical characteristics

We compared the clinical characteristics of the long COVID and control cohorts and reported odds ratios and 95% confidence intervals. For age, we considered the following buckets: <18, 18-24, 25-34, 35-44, 45-54, 55-64, 65-74, and 75+ years old. For race, we grouped the categories (“Asian,” “Asian - Far East,” and “Asian - Indian Subcontinent”) as “Asian,” and we grouped the categories (“Chose not to disclose,” “Unable to provide,” and “Unknown”) as “Unknown.” For ethnicity, we grouped the categories (“Choose not to disclose” and “Unknown”) as “Unknown.” For the number of lab test encounters within the past year, we considered the following buckets: 0, 1-3, and 4+ lab test encounters. Individuals with at least one dose of the Janssen COVID-19 vaccine or two or more doses of the Pfizer or Moderna COVID-19 vaccines on record were considered to be fully vaccinated. Comorbidities were determined based on ICD codes observed during the baseline phase. Comorbidities in the Charlson Comorbidity Index^14^ were considered along with auto-immune diseases and related conditions, including chronic fatigue syndrome, postural tachycardia syndrome without hypotension, fibromyalgia, and migraine. We also compared medications administered or ordered for the matched long COVID and control cohorts during the baseline, acute COVID-19, and post-COVID-19 phases. We report p-values from Fisher’s exact test performed for each phase.

### Statistical analysis software

All statistical analyses were performed using the Numpy (version 1.23.3), Scipy (version 1.9.1), and Statmodels (version 0.13.2) in Python 3.9.6.

## Results

The study population included 88,943 patients with a positive PCR test for SARS-CoV-2, including 1,140 patients with an ICD-10 code diagnosis for long COVID (U09.9). In **Table S3**, we provide the clinical characteristics of the unmatched cohorts. We observed that the observed rate of long COVID was higher among females compared to males (odds ratio: 1.42, 95% CI: [1.26, 1.60]). In addition, the median age of individuals in the long COVID cohort was significantly higher compared to the control cohort (**Table S3**). We performed a matched analysis to control for differences in demographics and other potential confounding factors for long COVID diagnosis (see **Methods** section for details). In **Table 1**, we provide the comorbidities and clinical outcomes for the final 1:1 matched long COVID and control cohorts. In **Table S4**, we provide a summary of the matched clinical characteristics for these cohorts. For the rest of this section, we present the results based on these matched cohorts.

**Table 1:** Comorbidities and clinical outcomes of long COVID and matched control cohorts. For each categorical variable, the percentage of patients in each cohort is shown along with the odds ratio and corresponding 95% confidence interval. Odds ratios that are statistically significant (p-value < 0.05) are indicated with *, and those that are highly significant (p-value < 0.001) are indicated with ***. Odds ratios for comparisons with <1% of patients in both cohorts are not shown. The matched clinical characteristics for these two cohorts are provided in **Table S4**.

|  | Long COVID cohort (matched) | Control cohort (matched) | Odds Ratio [95% CI] |
| --- | --- | --- | --- |
| **Number of individuals** | 1,086 | 1,086 | - |
| **Fully vaccinated before infection<sup>1</sup> (%)** |  |  |  |
| - Pfizer (two or more doses) | 37 | 41 | 0.85 [0.71, 1.00] |
| - Moderna (two or more doses) | 16 | 20 | 0.78 [0.62, 0.97]* |
| - Janssen (one or more doses) | 5 | 4 | 1.37 [0.90, 2.07] |
| - Any other vaccine (two or more doses) | 0 | <1 | - |
| **Charlson comorbidities in baseline phase (%)** |  |  |  |
| - Cancer | 6 | 6 | 0.98 [0.68, 1.41] |
| - Cerebrovascular disease | 3 | 3 | 0.94 [0.58, 1.53] |
| - Chronic pulmonary disease | 15 | 8 | 1.94 [1.48, 2.55]*** |
| - Congestive heart failure | 8 | 6 | 1.40 [1.00, 1.97] |
| - Dementia | <1 | <1 | - |
| - Diabetes without chronic complication | 12 | 10 | 1.14 [0.87, 1.50] |
| - Hemiplegia or paraplegia | <1 | <1 | 0.33 [0.07, 1.65] |
| - Metastatic solid tumor | 1 | 2 | 0.63 [0.34, 1.20] |
| - Mild liver disease | 3 | 5 | 0.69 [0.45, 1.06] |
| - Moderate or severe liver disease | <1 | <1 | - |
| - Myocardial infarction | 3 | 1 | 1.83 [0.99, 3.40] |
| - Peptic ulcer disease | 1 | <1 | 2.52 [0.97, 6.52] |
| - Peripheral vascular disease | 7 | 5 | 1.31 [0.92, 1.86] |
| - Renal disease | 14 | 10 | 1.40 [1.08, 1.82]* |
| - Rheumatic disease | 5 | 3 | 1.48 [0.97, 2.27] |
| - at least one of the listed comorbidities | 40 | 36 | 1.21 [1.01, 1.44]* |
| **Auto-immune diseases and potentially related conditions in baseline phase (%)** |  |  |  |
| - Chronic Fatigue Syndrome | 1 | <1 | 2.16 [0.88, 5.32] |
| - Postural Tachycardia Syndrome Without Hypotension | <1 | <1 | - |
| - Fibromyalgia | 4 | 2 | 2.25 [1.32, 3.84]* |
| - Migraine | 10 | 5 | 2.22 [1.57, 3.14]*** |
| - at least one of the listed conditions | 13 | 6 | 2.27 [1.67, 3.08]*** |
| **Individuals admitted 0-14 days post-infection (%)** |  |  |  |
| - Hospitalized | 5 | 1 | 4.74 [2.58, 8.70]*** |
| - ICU admission | 3 | 1 | 2.63 [1.34, 5.15]* |
| - Intubated | 3 | 1 | 2.29 [1.26, 4.16]* |

<sup>1</sup> Patients who have received COVID-19 vaccine doses from multiple manufacturers are also included here.

### Cough, difficulty breathing, and tiredness are the most commonly reported conditions for the long COVID cohort in the post-COVID-19 infection phase

Next, we compared the rates of phenotypes reported in the clinical notes for the long COVID and control cohorts. For each phenotype, we observed higher rates in the long COVID cohort compared to the control cohort during both the acute COVID-19 and post-COVID-19 phases (**Figure 1B, Figure S2**). Overall phenotype reporting was highest immediately following incidence of COVID-19, and for the long COVID cohort we found that phenotype reporting was increased compared to baseline reporting throughout (**Figure S2**). In contrast, phenotype reporting in the control cohort was back at baseline levels within 20 days of their positive PCR test (**Figure S2**). For the long COVID cohort, almost all of the phenotypes were reported at lower rates during the post-COVID-19 phase compared to the acute COVID-19 phase, with the exception of brain fog (increase from <1% to 3%) and sleep problems (increase from 2% to 3%), which were both higher during the post-COVID-19 phase. The most common phenotypes in the long COVID cohort during the post-COVID-19 phase were cough (14%), difficulty breathing (12%), and tiredness (10%).

### Comparison of patient characteristics before infection

To identify features associated with a higher risk of developing post-COVID-19 conditions, we assessed differences during the baseline phase. We observed that patients with chronic lung disease had higher rates of long COVID diagnosis (odds ratio: 1.94, 95% CI: [1.48, 2.55]) (**Table 1**). This subpopulation of patients with chronic lung disease included patients with asthma, COPD, emphysema, and bronchiectasis (**Figure S3**). We also observed that individuals with renal disease had higher rates of long COVID diagnosis (odds ratio: 1.40, 95% CI: [1.08, 1.82]). Auto-immune diseases and conditions including migraine (odds ratio: 2.40, 95% CI: [1.77, 3.25]) and fibromyalgia (odds ratio: 2.25, 95% CI: [1.32, 3.84]) were also more common as pre-existing conditions in the long COVID cohort.

### Comparison of lab test measurements during acute infection

To determine whether there are clinical signatures of acute COVID-19 disease indicative of increased risk for subsequent post-COVID-19 conditions, we assessed differences during the acute COVID-19 phase. We observed differences consistent with increased acute disease severity in the long COVID cohort compared with their matched controls. In the long COVID cohort, hospital admission rates (within 14 days of infection) were significantly increased (**Table 1**, rate_longCOVID_: 5% vs. rate_control_: 1%, p-value: <0.001, odds ratio: 4.74 [2.58, 8.70]). Similarly, ICU admission rates were also significantly higher in the long COVID cohort (rate_longCOVID_: 3% vs. rate_control_: 1%, p-value: <0.01, odds ratio: 2.63 [1.34, 5.15])).

To assess whether laboratory measurements could predict onset of long COVID, we analyzed measurements for 82 tests contributed by more than ten patients in both the long COVID and the control cohorts during acute SARS-CoV-2 infection (**Table S5**). For 15 lab tests, the long COVID cohort exhibited a significant difference in mean test results (p-value < 0.05) during the acute phase, both compared to the control cohort during the acute phase, and the long COVID cohort during the baseline phase. Further, we compared the test results for these 15 lab tests to their known normal ranges (shaded region, **Figure 1C** and **Figure S4, S5**) 6 out of these 15 tests in the long COVID cohort exhibited mean test results outside the normal range in the acute phase (**Figure 1C**). Specifically, we observed increased levels of: neutrophil-lymphocyte ratio (mean_longCOVID_: 10.7, 95% CI: [7.9, 14.3] vs. mean_control_: 7.2 [3.9, 11.0]), alanine aminotransferase (42.4 [38.0, 47.6] u/L vs. 36.7 [28.9, 46.4] u/L), and serum triglyceride (278.5 [203.5, 372.7] mg/dL vs. 141.4 [104.3, 187.0] mg/dL). We also observed decreased levels of serum HDL cholesterol (38.4 [32.9, 44.6] mg/dL vs. 52.5 [45.5, 60.3] mg/dL).

Concordant signals of more severe acute disease in long-COVID patients are also found when looking at medications ordered and administered during both the acute and post-acute phases (**Figure S6, Table S6**). Notably, antivirals, anticoagulants, and steroids were administered at significantly higher rates in the long COVID cohort. We did not observe a significant difference for monoclonal antibodies and administration of Albuterol was already elevated during the baseline, consistent with a higher prevalence of CPD in the long COVID cohort (**Figure S7, Table S6**).

## Discussion

In this study, we provide an in-depth characterization of a cohort of 1,086 patients diagnosed with long COVID compared with a matched control cohort. We found that the long COVID cohort was significantly enriched in patients with a history of CPD, fibromyalgia, and migraine. Additionally, we found that the patients that developed long COVID showed signs of more severe COVID-19 during their acute infection (0 to 14 days after infection) based on hospitalization, lab measurements, and medications administered.

Our findings are consistent with previous studies that have investigated long COVID signs and symptoms. We found that the most common phenotypes reported by long-COVID patients included cough, breathing difficulties, tiredness, and heart palpitations.^13,15^ We also found that long COVID patients exhibited low HDL cholesterol and high triglycerides levels in their serum during the acute COVID-19 phase, consistent with a previous retrospective study of 1,411 hospitalized COVID-19 patients.^16^ Previous studies have also shown that low albumin levels and elevated transaminases (ALT, AST) are associated with severe COVID-19 outcomes, and we observed the same to a lesser extent (**Figure S4**).^17–19^ Several of the long COVID characteristics observed in this study were also shown to be important variables for a recently described predictor of long COVID, including: difficulty breathing, dyspnea, cough, hospitalization, albuterol use, and CPD.^15^

In addition, both patients in the long COVID cohort and those in the control cohort had elevated levels of serum glucose level during the acute COVID-19 phase. In prior work, cases of metabolic dysfunction during and after SARS-CoV-2 infection have been reported ranging from new-onset diabetes mellitus (both Type I and Type 2) to asymptomatic insulin resistance and glucose intolerance.^20–23^ Acute SARS-CoV-2 infection can lead to metabolic dysfunction, including abnormal lipid profiles and sustained elevation in plasma glucose, through the chronic elaboration of cytokines, glucocorticoid treatment, and sustained stress related to severe infection and co-morbidity.^16,20,24^ There is evidence that these metabolic disturbances may be related to viral persistence in adipose tissue.^25^ Data from the current study also point to the potential for a metabolic signature. For example, changes in glucose disposal and lipid handling during the acute illness phase may be an early signal of further symptomatology and long COVID.

There are several limitations for this analysis. First, this is a retrospective study carried out in a single multi-state health system, so the clinical characteristics of the study population are not representative of the entire population of patients with post-COVID-19 conditions. Second, the ICD-10 code for post-COVID-19 conditions only became available in the United States on October 1, 2021,^26^ so this analysis was restricted to long COVID cases reported during the Delta and Omicron waves of the pandemic. Third, although we control for demographics, time and date of infection, and the number of prior lab tests, additional confounding factors might explain the differences in the long COVID and control cohorts. For example, individuals in the long COVID cohort may engage in more health-seeking behaviors and thus have higher rates of reported comorbidities than the control group. Additionally, since patients in the long COVID cohort were more likely hospitalized, their EHR data may be more complete. This may partially explain the observed higher rates of medications or disease symptoms in the long COVID cohort due to improved recording. We can therefore not draw causal relationships between observed enrichments and long COVID incidence. Fourth, not all of the patients in the long COVID and control cohorts underwent laboratory testing, so the observed distributions of lab values may not represent the distribution of lab values in the overall cohort. For example, some lab tests are ordered only in cases of a suspected diagnosis. In follow-up studies, methods for imputing missing values may be applied such as zero imputation, mean imputation, and multiple imputation.^27^

Overall, this study provides further clarity to the identification of risk factors for long COVID and motivates future research on the relationship between early interventions in COVID and the onset of long COVID. Future studies are needed to define individuals at highest risk for persistent symptomatology and possible interventions to forestall or prevent long COVID.

## Data Availability

After publication, the data will be made available to others upon reasonable requests to the corresponding author. A proposal with a detailed description of the study objectives and the statistical analysis plan will be needed for evaluation of the reasonability of requests.

## Supplementary Materials

**Figure S1:**
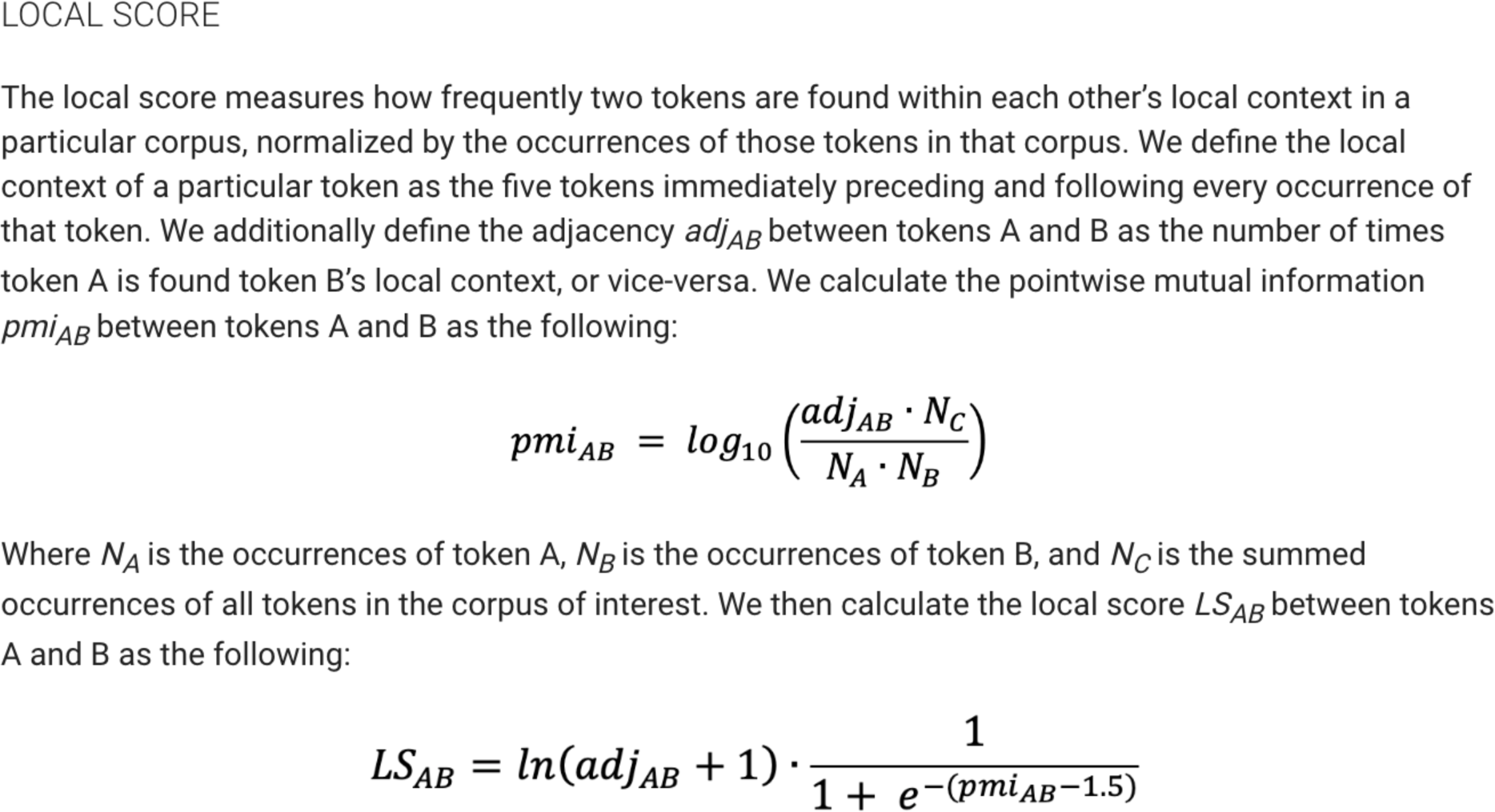
Formula for the nferX local score to measure literature associations.

**Figure S2:**
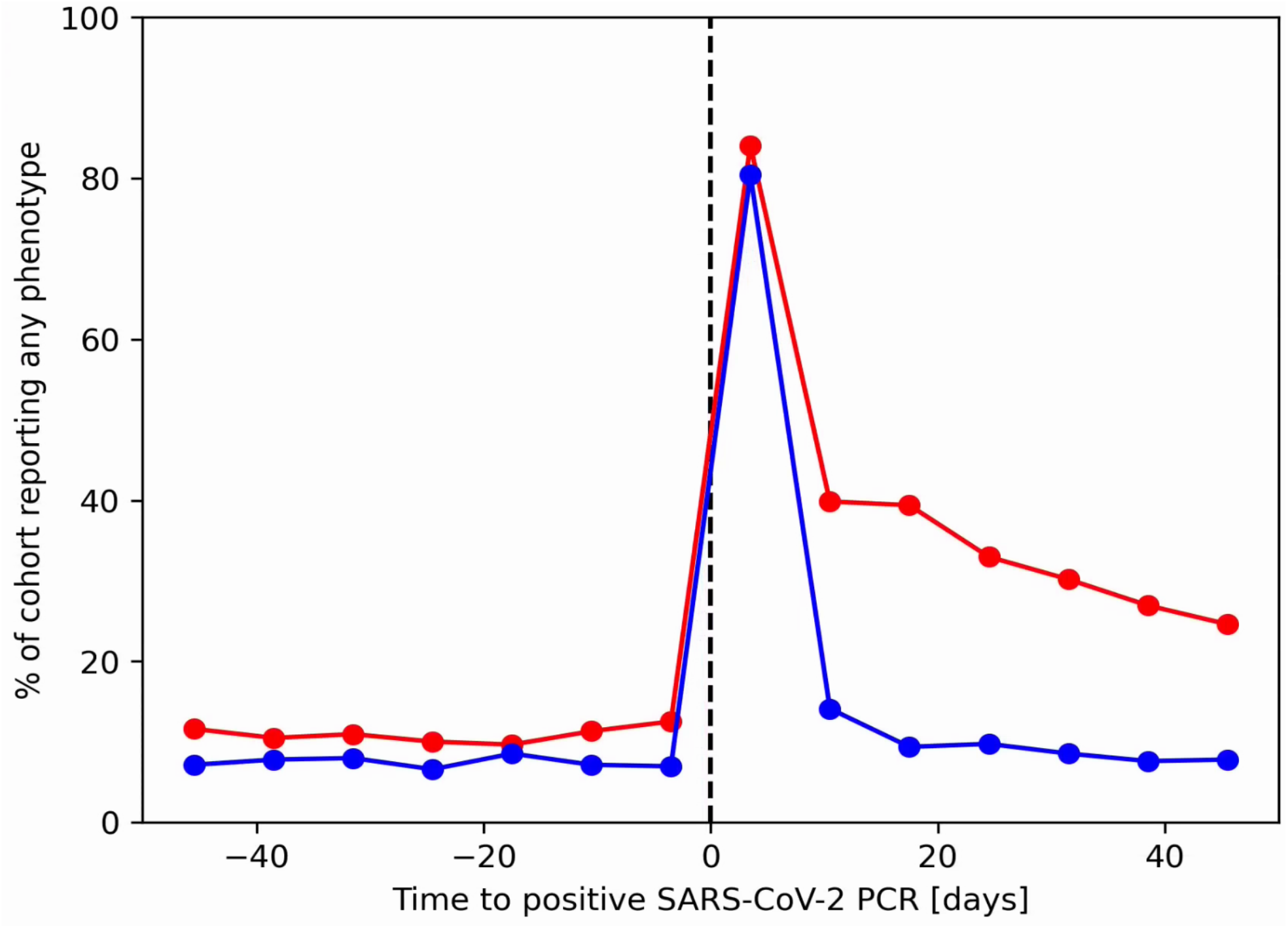
Reporting of any tracked phenotype as a function of time. Data shown for the long COVID cohort (red) and their 1:1 matched controls (blue). The vertical dashed line indicates the date of the patient’s positive SARS-CoV-2 PCR test.

**Figure S3:**
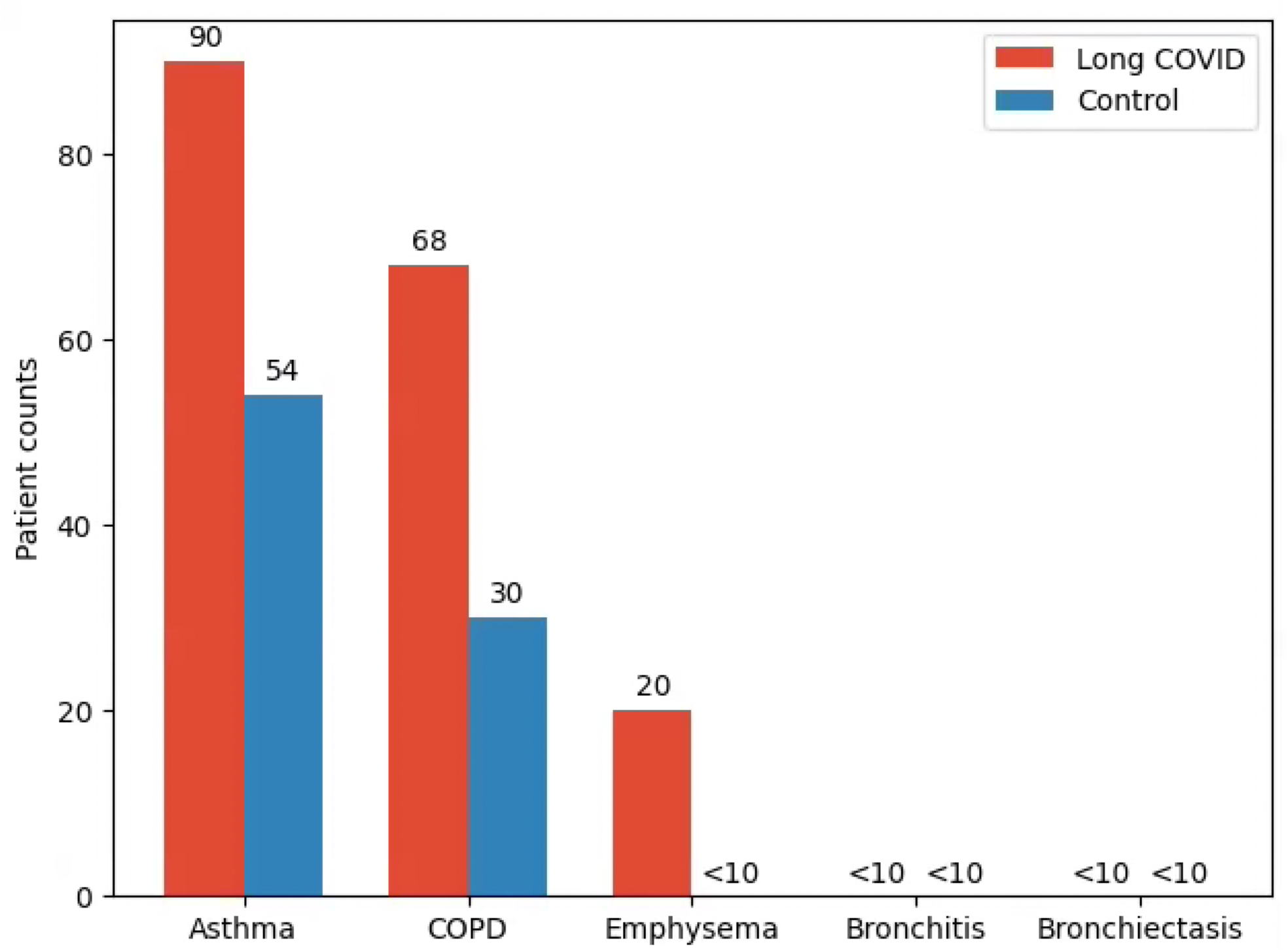
Subtypes of chronic pulmonary disorder in the long COVID and control cohorts.

**Figure S4:**
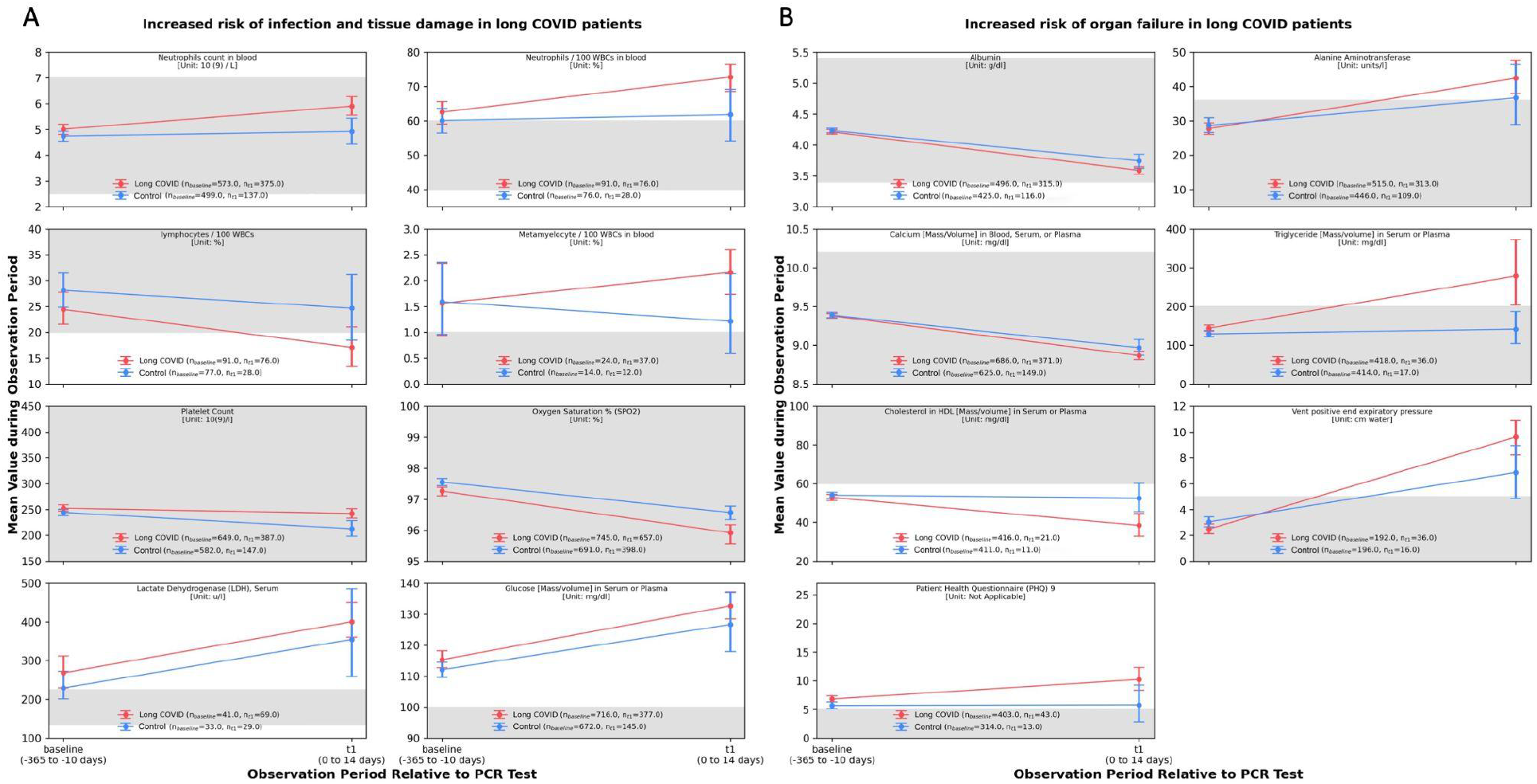
Lab test enrichments for matched long COVID and control cohorts. For each lab test, mean test values for the long COVID cohort were compared to those of the control cohort (see *Methods*). The error bars represent 95% confidence intervals, calculated by bootstrap resampling (1000 samples). The normal ranges for these lab tests^28–31^ are shaded in gray. Fifteen lab tests shown here are significantly different (Mann Whitney U test, p-value < 0.05) between the long COVID and the control cohorts in the acute COVID-19 phase and also significantly different between the long COVID cohorts in the baseline and acute COVID-19 phases. **(a)** Lab tests indicating infection and tissue damage **(b)** Lab tests indicating risk of organ failure.

**Figure S5:**
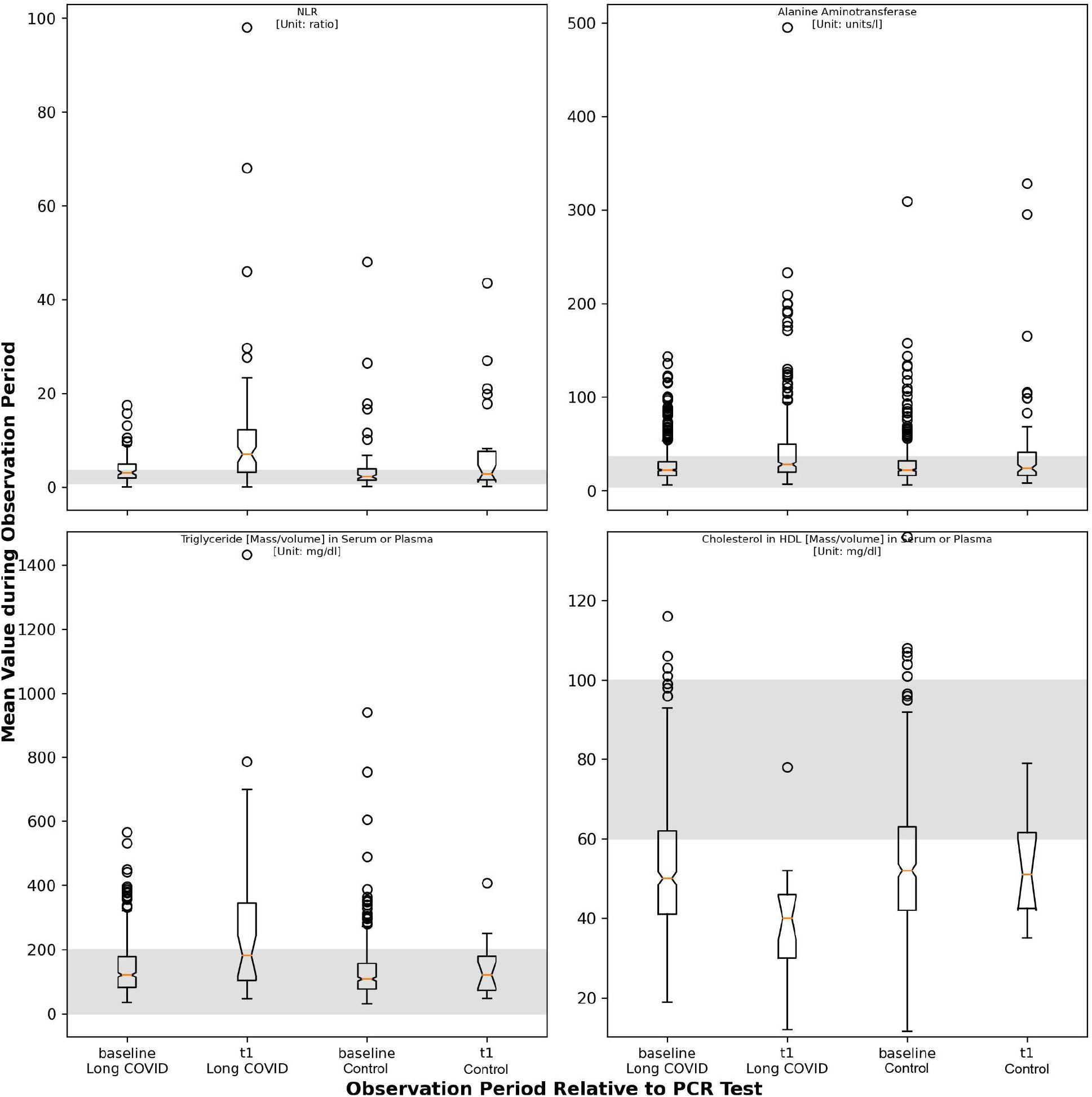
Lab test enrichments for matched long COVID and control cohorts. For each lab test, the distribution of mean_individual_ test values for the long COVID cohort were compared to those of the control cohort (see *Methods*). The error bars represent 95% confidence intervals, calculated by bootstrap resampling (1000 samples). The normal ranges for these lab tests^28–31^ are shaded in gray. Here we show four of these 15 lab tests significantly enriched in the long COVID cohort (see *Methods*) with mean test values outside the normal range.

**Figure S6:**
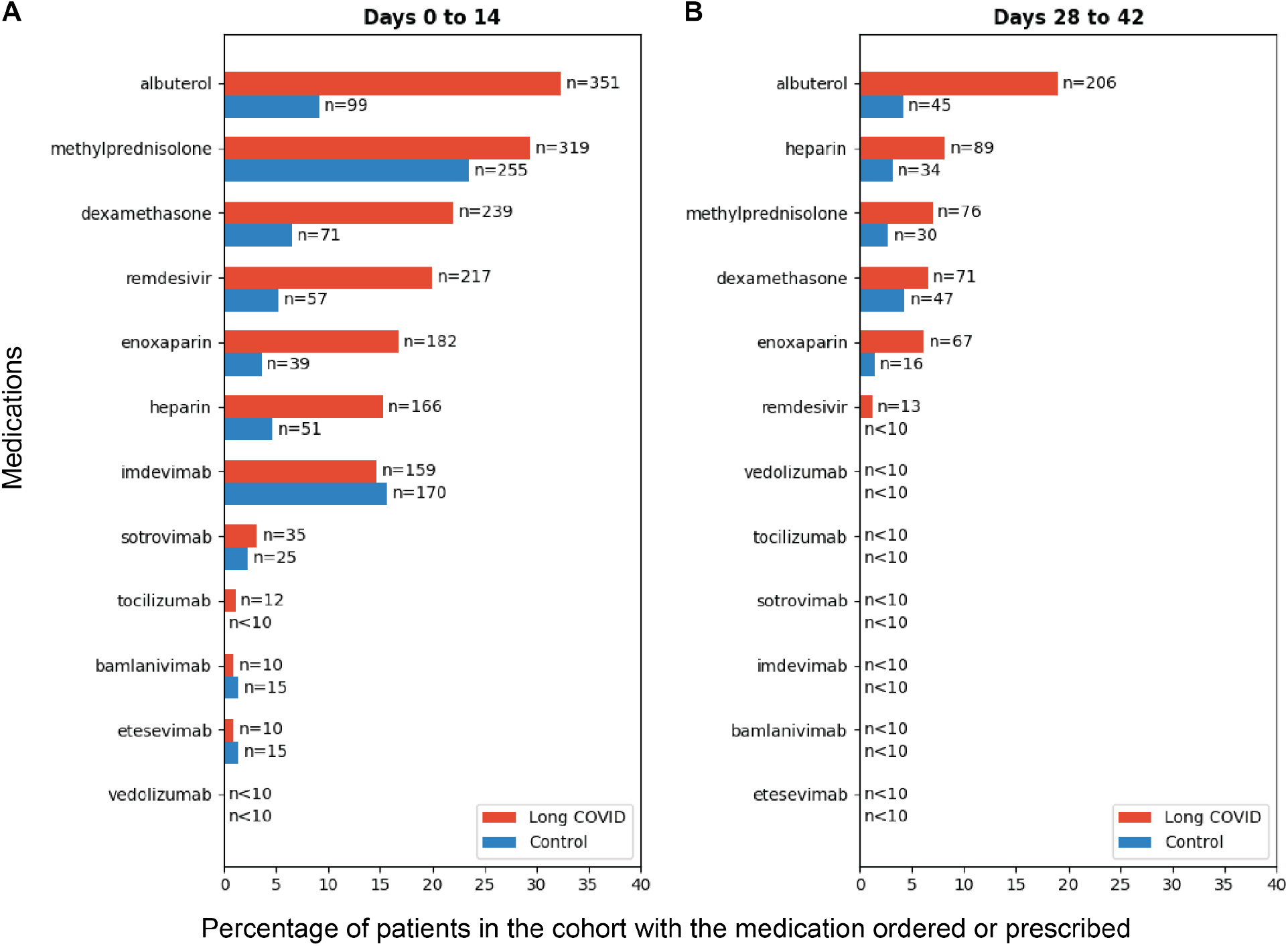
Comparison of medications administered or ordered for matched long COVID and control cohorts. Medications administered or ordered during the acute COVID-19 phase, **(A)**, the post-COVID-19 phase, **(B)** for the matched long COVID and control patients.

**Figure S7:**
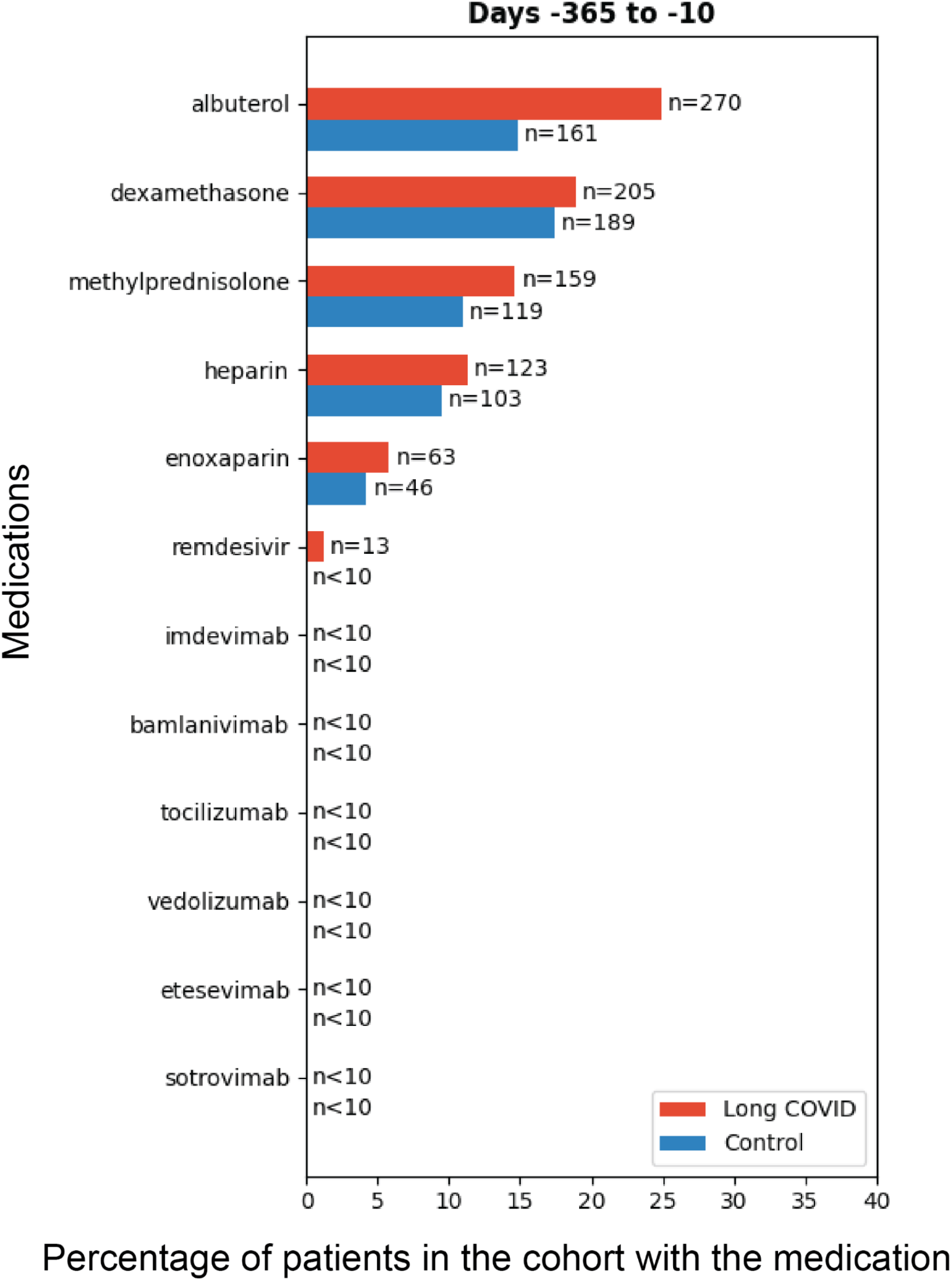
Comparison of medications administered or ordered for matched long COVID and control cohorts for the baseline phase.

**Table S1:** List of long COVID phenotypes identified by CDC and nferX Signals. In the first two columns, the phenotype names are shown along with the data source (e.g. CDC, Signals, or both). In the third column, the nferX Local Score is shown, which is a measure of the strength of the association between that phenotype and long COVID in the biomedical literature. Phenotypes with the highest local score values are most strongly associated with long COVID in the literature. (Link to nferX Signals query: https://nferx.com/turl/ODBlY2)

| Phenotype | Source | nferX Local Score |
| --- | --- | --- |
| brain_fog | CDC and Signals | 6.972 |
| anosmia | Signals | 5.155 |
| muscle_pain | CDC and Signals | 5.128 |
| severe_covid_19_disease | Signals | 5.121 |
| fibromyalgia | Signals | 5.054 |
| encephalomyelitis | Signals | 4.89 |
| chest_pain | CDC and Signals | 4.746 |
| dysautonomia | Signals | 4.729 |
| post_exertional_malaise | CDC and Signals | 4.683 |
| headache | CDC and Signals | 4.551 |
| acute_respiratory_syndrome | Signals | 4.531 |
| neurologic_manifestations | Signals | 4.527 |
| cognitive_dysfunction | Signals | 4.508 |
| cough | CDC and Signals | 4.372 |
| joint_pain | CDC and Signals | 4.323 |
| post_intensive_care_syndrome | CDC and Signals | 4.299 |
| orthostatic_intolerance | Signals | 4.257 |
| post_concussion_syndrome | Signals | 4.238 |
| pancreatic_adenosquamous_carcinoma | Signals | 4.199 |
| severe_acute_respiratory_syndrome | Signals | 4.122 |
| solid_tumor | Signals | 4.087 |
| anxiety | CDC and Signals | 4.059 |
| ageusia | Signals | 4.05 |
| myocarditis | Signals | 3.998 |
| dysgeusia | CDC and Signals | 3.959 |
| adenosquamous_carcinoma | Signals | 3.945 |
| pacs | Signals | 3.911 |
| cocaine_intoxication | Signals | 3.897 |
| pulmonary_fibrosis | Signals | 3.827 |
| muscle_weakness | CDC and Signals | 3.741 |
| tiredness | CDC and Signals | 3.634 |
| immune_dysregulation | Signals | 3.615 |
| parosmia | CDC and Signals | 3.601 |
| postural_orthostatic_tachycardia_syndrome | Signals | 3.599 |
| pancreatic_ductal_adenocarcinoma | Signals | 3.56 |
| depression | CDC and Signals | 3.548 |
| mast_cell_activation_syndrome | Signals | 3.521 |
| non_alcoholic_steatohepatitis | Signals | 3.479 |
| alcoholic_steatohepatitis | Signals | 3.415 |
| body_aches | Signals | 3.395 |
| viral_infection | Signals | 3.366 |
| post_infectious_syndromes | Signals | 3.315 |
| inappropriate_sinus_tachycardia | Signals | 3.298 |
| fever | CDC and Signals | 3.189 |
| poisoning | Signals | 3.181 |
| cognitive_deficits | Signals | 3.179 |
| acute_respiratory_distress_syndrome | Signals | 3.178 |
| lung_disease | Signals | 3.127 |
| hepatic_encephalopathy | Signals | 3.075 |
| secondary_lymphedema | Signals | 3.009 |
| autoimmune_conditions | CDC | < 3 |
| changes_in_menstrual_cycles | CDC | < 3 |
| diabete_mellitus | CDC | < 3 |
| diarrhea | CDC | < 3 |
| difficulty_breathing | CDC | < 3 |
| heart_palpitations | CDC | < 3 |
| lightheadedness | CDC | < 3 |
| multisystem_inflammatory_syndrome | CDC | < 3 |
| myalgic_encephalomyelitis | CDC | < 3 |
| pins_and_needles_feelings | CDC | < 3 |
| post_traumatic_stress_disorder | CDC | < 3 |
| rash | CDC | < 3 |
| sleep_problems | CDC | < 3 |
| stomach_pain | CDC | < 3 |

**Table S2:**
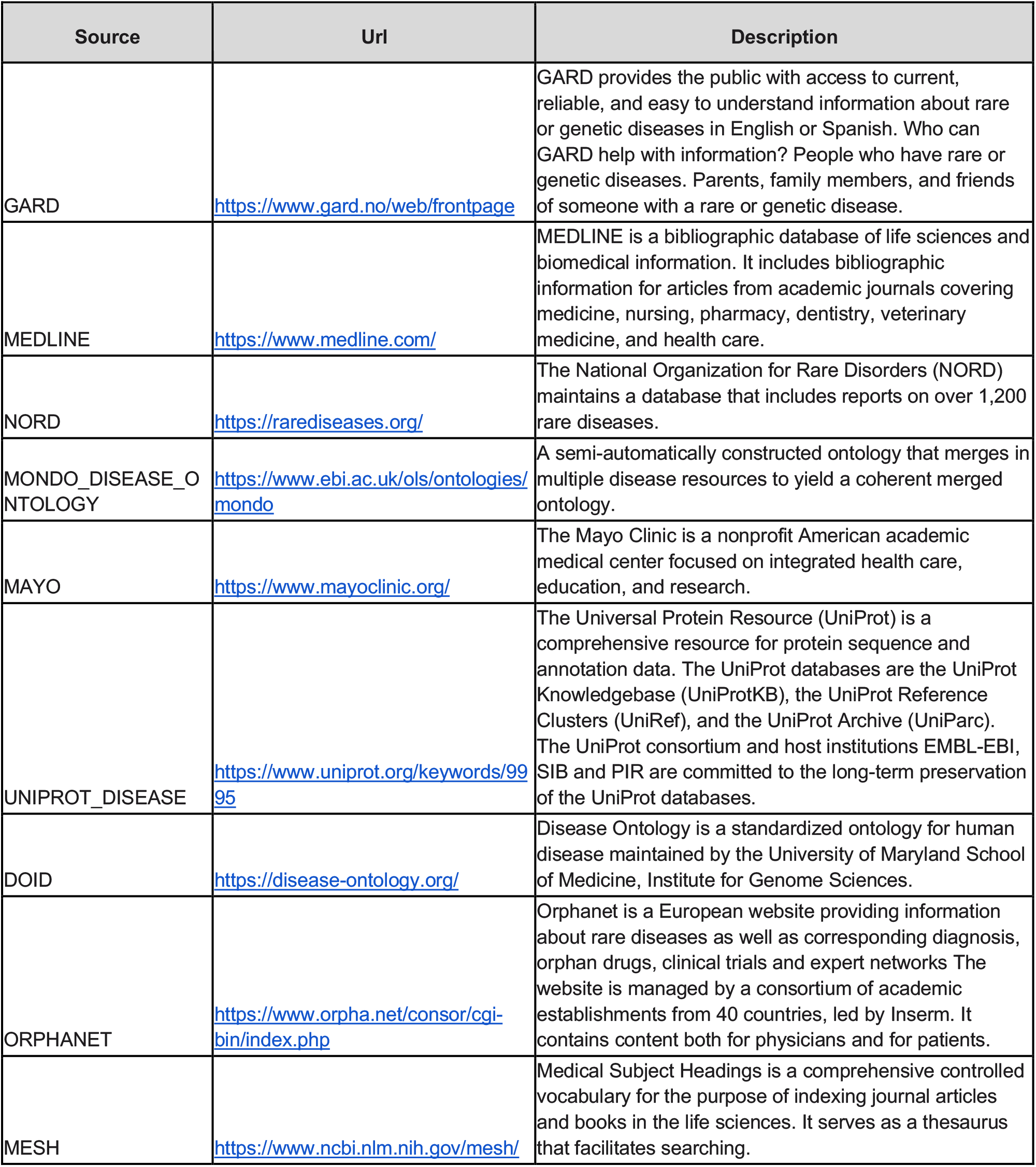
List of data sources for nferX Diseases collection.

**Table S3:**
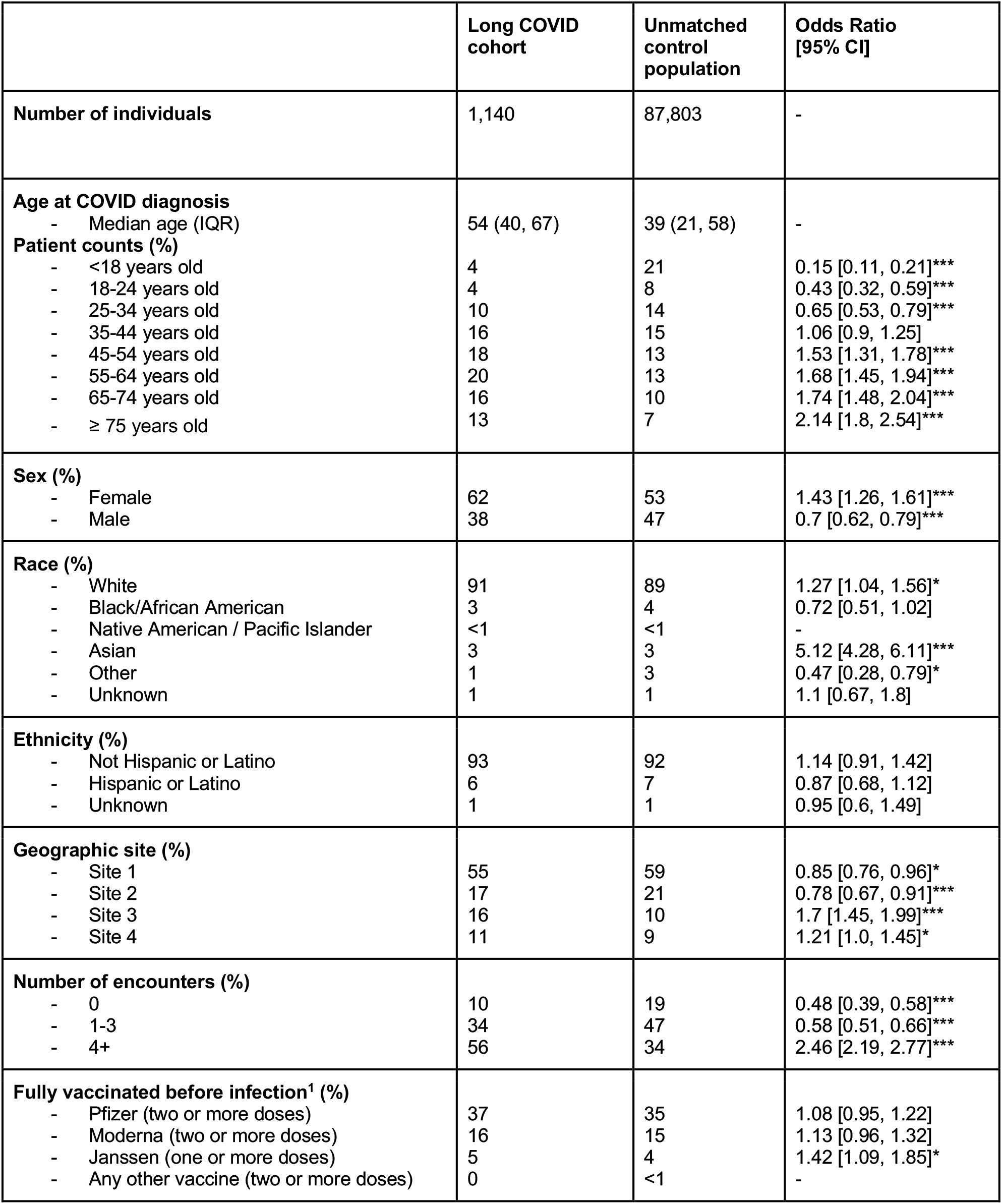
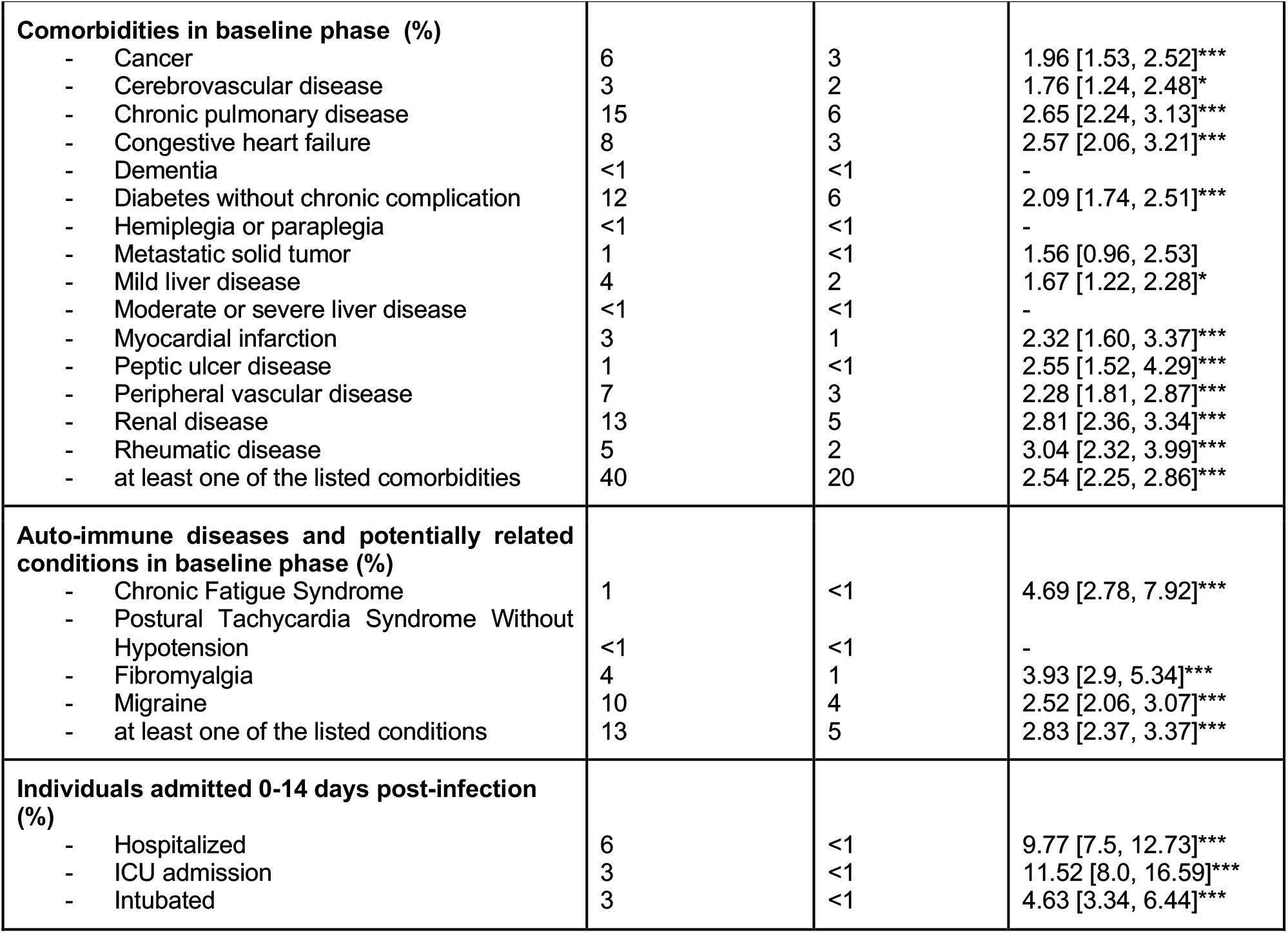
Clinical characteristics, comorbidities, and clinical outcomes of long COVID and pre-matching control population. For each categorical variable, the percentage of patients in each cohort is shown along with the odds ratio and corresponding 95% confidence interval. Odds ratios that are statistically significant (p-value < 0.05) are indicated with *, and those that are highly significant (p-value < 0.001) are indicated with ***.

|  | Long COVID cohort | Unmatched control population | Odds Ratio [95% CI] |
| --- | --- | --- | --- |
| **Number of individuals** | 1,140 | 87,803 | - |
| **Age at COVID diagnosis**<br>- Median age (IQR) | 54 (40, 67) | 39 (21, 58) | - |
| **Patient counts (%)**<br>- <18 years old | 4 | 21 | 0.15 [0.11, 0.21]*** |
| - 18-24 years old | 4 | 8 | 0.43 [0.32, 0.59]*** |
| - 25-34 years old | 10 | 14 | 0.65 [0.53, 0.79]*** |
| - 35-44 years old | 16 | 15 | 1.06 [0.9, 1.25] |
| - 45-54 years old | 18 | 13 | 1.53 [1.31, 1.78]*** |
| - 55-64 years old | 20 | 13 | 1.68 [1.45, 1.94]*** |
| - 65-74 years old | 16 | 10 | 1.74 [1.48, 2.04]*** |
| - ≥ 75 years old | 13 | 7 | 2.14 [1.8, 2.54]*** |
| **Sex (%)**<br>- Female | 62 | 53 | 1.43 [1.26, 1.61]*** |
| - Male | 38 | 47 | 0.7 [0.62, 0.79]*** |
| **Race (%)**<br>- White | 91 | 89 | 1.27 [1.04, 1.56]* |
| - Black/African American | 3 | 4 | 0.72 [0.51, 1.02] |
| - Native American / Pacific Islander | <1 | <1 | - |
| - Asian | 3 | 3 | 5.12 [4.28, 6.11]*** |
| - Other | 1 | 3 | 0.47 [0.28, 0.79]* |
| - Unknown | 1 | 1 | 1.1 [0.67, 1.8] |
| **Ethnicity (%)**<br>- Not Hispanic or Latino | 93 | 92 | 1.14 [0.91, 1.42] |
| - Hispanic or Latino | 6 | 7 | 0.87 [0.68, 1.12] |
| - Unknown | 1 | 1 | 0.95 [0.6, 1.49] |
| **Geographic site (%)**<br>- Site 1 | 55 | 59 | 0.85 [0.76, 0.96]* |
| - Site 2 | 17 | 21 | 0.78 [0.67, 0.91]*** |
| - Site 3 | 16 | 10 | 1.7 [1.45, 1.99]*** |
| - Site 4 | 11 | 9 | 1.21 [1.0, 1.45]* |
| **Number of encounters (%)**<br>- 0 | 10 | 19 | 0.48 [0.39, 0.58]*** |
| - 1-3 | 34 | 47 | 0.58 [0.51, 0.66]*** |
| - 4+ | 56 | 34 | 2.46 [2.19, 2.77]*** |
| **Fully vaccinated before infection<sup>1</sup> (%)**<br>- Pfizer (two or more doses) | 37 | 35 | 1.08 [0.95, 1.22] |
| - Moderna (two or more doses) | 16 | 15 | 1.13 [0.96, 1.32] |
| - Janssen (one or more doses) | 5 | 4 | 1.42 [1.09, 1.85]* |
| - Any other vaccine (two or more doses) | 0 | <1 | - |
| **Comorbidities in baseline phase (%)** |  |  |  |
| - Cancer | 6 | 3 | 1.96 [1.53, 2.52]*** |
| - Cerebrovascular disease | 3 | 2 | 1.76 [1.24, 2.48]* |
| - Chronic pulmonary disease | 15 | 6 | 2.65 [2.24, 3.13]*** |
| - Congestive heart failure | 8 | 3 | 2.57 [2.06, 3.21]*** |
| - Dementia | <1 | <1 | - |
| - Diabetes without chronic complication | 12 | 6 | 2.09 [1.74, 2.51]*** |
| - Hemiplegia or paraplegia | <1 | <1 | - |
| - Metastatic solid tumor | 1 | <1 | 1.56 [0.96, 2.53] |
| - Mild liver disease | 4 | 2 | 1.67 [1.22, 2.28]* |
| - Moderate or severe liver disease | <1 | <1 | - |
| - Myocardial infarction | 3 | 1 | 2.32 [1.60, 3.37]*** |
| - Peptic ulcer disease | 1 | <1 | 2.55 [1.52, 4.29]*** |
| - Peripheral vascular disease | 7 | 3 | 2.28 [1.81, 2.87]*** |
| - Renal disease | 13 | 5 | 2.81 [2.36, 3.34]*** |
| - Rheumatic disease | 5 | 2 | 3.04 [2.32, 3.99]*** |
| - at least one of the listed comorbidities | 40 | 20 | 2.54 [2.25, 2.86]*** |
| **Auto-immune diseases and potentially related conditions in baseline phase (%)** |  |  |  |
| - Chronic Fatigue Syndrome | 1 | <1 | 4.69 [2.78, 7.92]*** |
| - Postural Tachycardia Syndrome Without Hypotension | <1 | <1 | - |
| - Fibromyalgia | 4 | 1 | 3.93 [2.9, 5.34]*** |
| - Migraine | 10 | 4 | 2.52 [2.06, 3.07]*** |
| - at least one of the listed conditions | 13 | 5 | 2.83 [2.37, 3.37]*** |
| **Individuals admitted 0-14 days post-infection (%)** |  |  |  |
| - Hospitalized | 6 | <1 | 9.77 [7.5, 12.73]*** |
| - ICU admission | 3 | <1 | 11.52 [8.0, 16.59]*** |
| - Intubated | 3 | <1 | 4.63 [3.34, 6.44]*** |

**Table S4:** Clinical characteristics of long COVID and matched control cohorts. For each categorical variable, the percentage of patients in each cohort is shown. During the matching procedure, each of the categorical variables were matched exactly, so the distributions are exactly the same for the two cohorts. The numeric variables (age and number of encounters) were bucket matched, so there may be slight differences in these covariates between the two cohorts.

|  | Long COVID cohort<br>(matched) | Control cohort<br>(matched) |
| --- | --- | --- |
| **Number of individuals** | 1,086 | 1,086 |
| **Age at COVID diagnosis (in years)**<br>- Median age (IQR) | 54 (41 - 68) | 54 (40 - 67) |
| **Age at COVID diagnosis (distribution)**<br>- <18 years old<br>- 18-24 years old<br>- 25-34 years old<br>- 35-44 years old<br>- 45-54 years old<br>- 55-64 years old<br>- 65-74 years old<br>- ≥ 75 years old | 4<br>4<br>9<br>16<br>18<br>19<br>16<br>14 | 4<br>4<br>9<br>16<br>18<br>19<br>16<br>14 |
| **Sex (%)**<br>- Female<br>- Male | 63<br>37 | 63<br>37 |
| **Race (%)**<br>- White<br>- Black/African American<br>- Native American / Pacific Islander<br>- Asian<br>- Other<br>- Unknown | 94<br>3<br><1<br>2<br><1<br><1 | 94<br>3<br><1<br>2<br>1<br>0 |
| **Ethnicity (%)**<br>- Not Hispanic or Latino<br>- Hispanic or Latino<br>- Unknown | 95<br>5<br><1 | 95<br>5<br><1 |
| **Geographic site (%)**<br>- Site 1<br>- Site 2<br>- Site 3<br>- Site 4 | 57<br>17<br>16<br>11 | 57<br>17<br>16<br>11 |
| **Number of encounters (%)**<br>- 0<br>- 1-3<br>- 4+ | 10<br>34<br>57 | 10<br>34<br>57 |

**Table S5:** List of lab tests in the acute COVID-19 phase. This list includes 82 lab tests and measurements contributed by more than 10 patients in both the long COVID and the control cohorts in the acute COVID-19 phase and is considered for further downstream analyses.

| Lab test/measurement | Unit | Long COVID |  |  | Control |  |  |
| --- | --- | --- | --- | --- | --- | --- | --- |
|  |  | Patient count | Mean | 95% CI | Patient count | Mean | 95% CI |
| Alanine Aminotransferase | units/l | 313 | 42.42 | [38.02, 47.63] | 109 | 36.7 | [28.93, 46.36] |
| Albumin | g/dl | 315 | 3.59 | [3.53, 3.65] | 116 | 3.74 | [3.62, 3.85] |
| Alkaline Phosphatase, Serum | U/L | 310 | 88.36 | [83.6, 93.68] | 110 | 92.53 | [83.39, 103.99] |
| anion gap, serum/plasma | not applicable | 372 | 11.58 | [11.36, 11.81] | 141 | 11.45 | [11.09, 11.83] |
| Aspartate Aminotransferase | unit/l | 312 | 48.41 | [40.31, 61.58] | 112 | 43.36 | [35.71, 54.81] |
| base excess arterial blood | mEq/L | 66 | 0.39 | [-0.55, 1.29] | 16 | -0.25 | [-2.26, 1.71] |
| Basophils | 10 (9)/l | 304 | 0.03 | [0.03, 0.04] | 106 | 0.03 | [0.03, 0.04] |
| Basophils / 100 WBCs | % | 37 | 0.55 | [0.36, 0.75] | 19 | 0.42 | [0.27, 0.59] |
| Bicarbonate, Arterial (HCO<sub>3</sub>) | mmol/L | 69 | 24.21 | [23.32, 25.17] | 17 | 23.28 | [21.49, 25.21] |
| Bicarbonate, Venous (HCO<sub>3</sub>) | mmol/L | 176 | 24.48 | [24.0, 24.93] | 62 | 24.74 | [23.9, 25.57] |
| Body Mass Index (BMI) | kg/m<sup>2</sup> | 332 | 16.05 | [14.92, 17.15] | 134 | 15.93 | [14.59, 17.36] |
| C Reactive Protein, in Serum/Plasma/Blood | mg/l | 239 | 63.18 | [55.69, 71.73] | 66 | 50.92 | [41.97, 60.82] |
| Calcium [Mass/Volume] in Blood, Serum, or Plasma | mg/dl | 371 | 8.87 | [8.81, 8.92] | 149 | 8.97 | [8.87, 9.07] |
| Calcium, Ionized, Serum | mg/dl | 59 | 4.73 | [4.66, 4.8] | 18 | 4.8 | [4.59, 5.07] |
|  |  | Patient count | Mean | 95% CI | Patient count | Mean | 95% CI |
| Carbon Dioxide [Partial Pressure], Arterial Blood (PaCO<sub>2</sub>) | mmhg | 69 | 37.36 | [35.51, 39.24] | 17 | 38.83 | [35.26, 42.12] |
| carboxyhaemoglobin | % | 68 | 0.94 | [0.8, 1.14] | 16 | 0.7 | [0.48, 0.98] |
| carboxyhaemoglobin (fractional) | % | 54 | 0.83 | [0.73, 0.92] | 13 | 0.66 | [0.44, 0.97] |
| Carboxyhemoglobin (Fractional), Arterial | % | 54 | 0.83 | [0.73, 0.92] | 13 | 0.66 | [0.42, 0.95] |
| Chloride [Moles/volume] in Blood, Plasma or Serum | mmol/l | 374 | 101.43 | [101.07, 101.83] | 142 | 101.78 | [101.15, 102.41] |
| Cholesterol in HDL [Mass/volume] in Serum or Plasma | mg/dl | 21 | 38.43 | [32.86, 44.62] | 11 | 52.55 | [45.55, 60.28] |
| Creatinine | mg/dl | 380 | 1.06 | [0.99, 1.13] | 150 | 1.17 | [1.03, 1.34] |
| D - dimer | ng/ml feu | 272 | 2013.63 | [1665.64, 2398.06] | 71 | 1917.63 | [1354.12, 2543.8] |
| Direct Bilirubin | mg/dL | 86 | 0.4 | [0.28, 0.57] | 29 | 0.37 | [0.24, 0.54] |
| Eosinophils [# / Volume], Blood | 10(9)/l | 325 | 0.1 | [0.09, 0.12] | 118 | 0.1 | [0.08, 0.12] |
| Eosinophils / 100 WBCs in blood | % | 49 | 1.51 | [1.09, 1.99] | 21 | 1.23 | [0.75, 1.85] |
| Erythrocyte Count | x10(12) /l | 386 | 4.29 | [4.22, 4.35] | 146 | 4.26 | [4.15, 4.38] |
| Erythrocyte Distribution Width (RDW), Blood | ratio | 386 | 13.91 | [13.73, 14.11] | 146 | 13.87 | [13.59, 14.2] |
| Erythrocyte Distribution Width (RDW), Entic Volume, Blood | fL | 30 | 47.34 | [43.91, 51.48] | 18 | 46.48 | [43.13, 50.52] |
| Estimated Glomerular Filtration Rate (eGFR) -- CKD-EPI | mL/min/BSA | 258 | 66.11 | [63.71, 68.5] | 96 | 62.73 | [59.22, 66.48] |
|  |  | Patient count | Mean | 95% CI | Patient count | Mean | 95% CI |
| Ferritin | mcg/l | 65 | 742.19 | [567.55, 913.99] | 21 | 723.72 | [426.73, 1085.46] |
| Fibrinogen [Mass/volume], Plasma | mg/dl | 75 | 506.85 | [473.64, 543.96] | 28 | 503.77 | [428.52, 578.28] |
| fractional oxyhemoglobin, blood | % | 53 | 82.7 | [77.59, 87.67] | 14 | 75.69 | [66.65, 84.83] |
| Glucose [Mass/volume] in Serum or Plasma | mg/dl | 377 | 132.74 | [128.48, 137.22] | 145 | 126.7 | [117.92, 137.05] |
| Heart Rate | bpm | 329 | 81.56 | [80.25, 83.01] | 112 | 82.06 | [79.66, 84.5] |
| Height | Meters | 263 | 1.67 | [1.65, 1.68] | 106 | 1.65 | [1.61, 1.68] |
| Hematocrit | % | 386 | 38.5 | [37.98, 39.06] | 147 | 38.42 | [37.56, 39.34] |
| Hemoglobin | g/dl | 387 | 12.75 | [12.56, 12.93] | 148 | 12.72 | [12.36, 13.04] |
| Immature granulocytes / 100 WBCs | % | 24 | 1.4 | [0.98, 1.84] | 12 | 0.93 | [0.61, 1.25] |
| Lactate | mmol/l | 42 | 1.59 | [1.26, 2.06] | 16 | 1.59 | [1.22, 2.12] |
| Lactate Dehydrogenase (LDH), Serum | u/l | 69 | 400.69 | [360.17, 450.1] | 29 | 354.68 | [259.51, 486.24] |
| LDL Cholesterol [Mass/volume] in Serum or Plasma | mg/dl | 19 | 80.84 | [67.84, 95.21] | 11 | 86.73 | [72.18, 101.09] |
| Leukocyte Count | 10(9)/l | 387 | 8.61 | [7.76, 9.95] | 147 | 8.08 | [6.93, 9.54] |
| Lipase, Serum | U/L | 46 | 46.97 | [37.32, 58.51] | 16 | 95.93 | [41.93, 189.58] |
| lymphocytes / 100 WBCs | % | 76 | 17.07 | [13.44, 21.02] | 28 | 24.65 | [18.45, 31.16] |
| Lymphocytes concentration in blood | 10(9)/l | 371 | 1.95 | [1.24, 3.13] | 136 | 2.38 | [1.43, 3.9] |
|  |  | Patient count | Mean | 95% CI | Patient count | Mean | 95% CI |
| Magnesium [Mass/Volume], in Serum, Plasma or Blood | mg/dl | 202 | 2.07 | [2.03, 2.11] | 59 | 2.03 | [1.97, 2.09] |
| Mean Airway Pressure | cm water | 37 | 13.04 | [11.14, 14.84] | 14 | 11.04 | [8.09, 13.95] |
| Mean arterial pressure | mmHg | 375 | 91.26 | [90.2, 92.3] | 156 | 90.49 | [88.93, 92.21] |
| Mean Corpuscular Hemoglobin (MCH) | pg | 30 | 29.31 | [28.26, 30.43] | 18 | 30.32 | [29.55, 31.08] |
| Mean Corpuscular Hemoglobin Concentration | g/dl | 30 | 32.56 | [31.92, 33.17] | 18 | 32.76 | [32.31, 33.22] |
| Mean Corpuscular Volume (MCV), Blood | fL | 386 | 90.19 | [89.67, 90.77] | 146 | 90.93 | [89.82, 92.02] |
| Mean Platelet Volume | fl | 30 | 10.44 | [10.12, 10.76] | 18 | 10.17 | [9.87, 10.48] |
| Metamyelocyte / 100 WBCs in blood | % | 37 | 2.16 | [1.73, 2.6] | 12 | 1.21 | [0.59, 2.13] |
| Monocytes [# / Volume] in Blood | 10(9)/l | 368 | 0.63 | [0.6, 0.66] | 134 | 0.66 | [0.58, 0.79] |
| Monocytes / 100 WBCs in blood | % | 75 | 7.04 | [6.08, 8.07] | 27 | 10.15 | [7.04, 14.21] |
| Myelocytes / 100 WBCs in blood | % | 29 | 2.78 | [1.91, 3.8] | 13 | 2 | [1.03, 3.33] |
| Natriuretic peptide B prohormone N-terminal | pg/ml | 117 | 2020.98 | [1321.17, 2830.31] | 27 | 1656.09 | [842.14, 2635.96] |
| Neutrophils / 100 WBCs in blood | % | 76 | 72.73 | [68.61, 76.42] | 28 | 61.81 | [54.14, 69.15] |
| Neutrophils count in blood | 10 (9) / L | 375 | 5.91 | [5.55, 6.27] | 137 | 4.93 | [4.43, 5.44] |
| NLR | ratio | 76 | 10.66 | [7.91, 14.33] | 28 | 7.19 | [3.93, 11.07] |
|  |  | Patient count | Mean | 95% CI | Patient count | Mean | 95% CI |
| Oxygen [Partial Pressure], Arterial Blood (PaO<sub>2</sub>) | mmHg | 69 | 82.38 | [76.89, 88.65] | 17 | 109.27 | [82.0, 146.69] |
| Oxygen [Partial Pressure], Venous Blood (PvO<sub>2</sub>) | mmHg | 74 | 36.39 | [33.66, 39.55] | 16 | 31.39 | [27.69, 35.33] |
| Oxygen Saturation % (SPO<sub>2</sub>) | % | 657 | 95.91 | [95.56, 96.17] | 398 | 96.57 | [96.34, 96.78] |
| Oxyhemoglobin (Fractional), Arterial | % | 48 | 92.83 | [92.19, 93.47] | 11 | 92.14 | [89.67, 94.03] |
| Patient Health Questionnaire (PHQ) 9 | Not Applicable | 43 | 10.23 | [8.32, 12.3] | 13 | 5.77 | [2.84, 9.23] |
| pH, U | pH | 69 | 5.73 | [5.6, 5.88] | 35 | 5.91 | [5.72, 6.12] |
| pH, venous blood | Not Applicable | 75 | 7.4 | [7.38, 7.41] | 17 | 7.39 | [7.37, 7.41] |
| Phosphate [Mass/volume] in Serum, Plasma or Blood | mg/dl | 78 | 3.55 | [3.3, 3.82] | 28 | 3.41 | [3.13, 3.7] |
| Platelet Count | 10<sup>9</sup>/l | 387 | 242.17 | [233.1, 251.54] | 147 | 212.56 | [198.06, 228.05] |
| Potassium [Moles/volume] in Serum or Plasma | mmol/l | 377 | 4.13 | [4.09, 4.18] | 145 | 4.13 | [4.05, 4.2] |
| Procalcitonin | ng/ml | 78 | 0.9 | [0.37, 1.79] | 17 | 0.44 | [0.19, 0.82] |
| Prothrombin time | seconds | 147 | 17.23 | [15.6, 18.91] | 48 | 16.24 | [13.84, 19.36] |
| Sodium [Moles / Volume] in Serum, Plasma or Blood | mmol/l | 377 | 137.34 | [137.0, 137.63] | 145 | 137.63 | [136.99, 138.2] |
| Total Bilirubin | mg/dl | 302 | 0.53 | [0.48, 0.6] | 102 | 0.59 | [0.51, 0.7] |
| Total Cholesterol [Mass/volume] in Serum or Plasma | mg/dl | 22 | 144.41 | [128.4, 161.41] | 12 | 157.25 | [140.5, 175.5] |
|  |  | Patient count | Mean | 95% CI | Patient count | Mean | 95% CI |
| Total Protein | g/dl | 311 | 6.42 | [6.34, 6.5] | 107 | 6.53 | [6.39, 6.69] |
| Triglyceride [Mass/volume] in Serum or Plasma | mg/dl | 36 | 278.52 | [203.5, 372.65] | 17 | 141.43 | [104.29, 187.0] |
| Troponin T, Cardiac [Mass/volume] | ng/ml | 165 | 0.03 | [0.03, 0.04] | 49 | 0.04 | [0.03, 0.06] |
| Urine Output | mL | 222 | 331.21 | [308.84, 352.76] | 58 | 268.16 | [234.29, 306.51] |
| urobilinogen, urine | mg/dL | 64 | 0.6 | [0.48, 0.74] | 30 | 0.69 | [0.45, 1.01] |
| Vent positive end expiratory pressure | cm water | 36 | 9.61 | [8.23, 10.88] | 16 | 6.87 | [4.85, 8.93] |
| Weight | Kg | 100 | 72.57 | [69.92, 75.19] | 25 | 69.15 | [64.51, 74.07] |

**Table S6:**
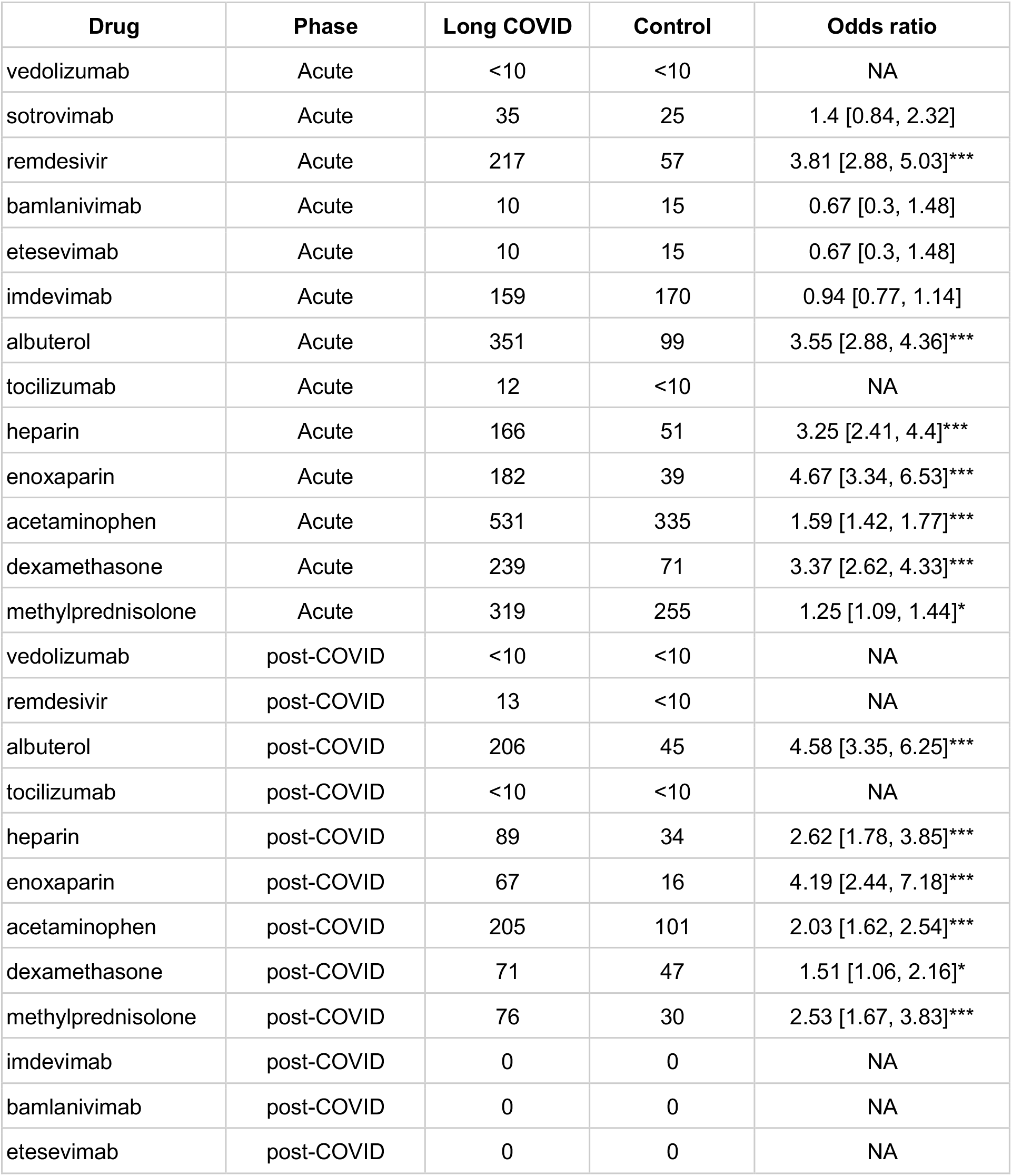
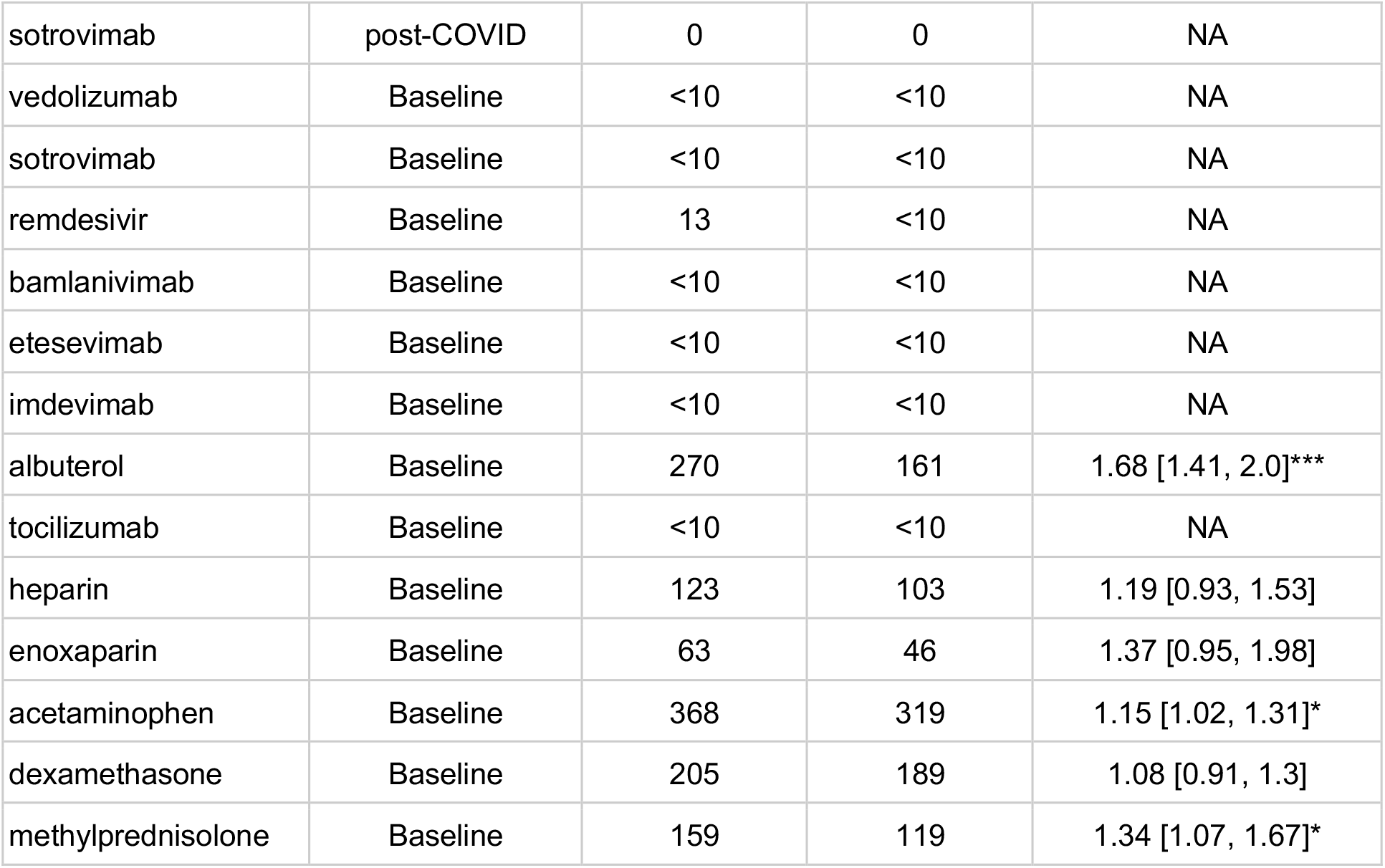
Comparison of medications administered or ordered during the acute, post COVID-19 and baseline phases. For each drug, the number of patients in each cohort is shown along with the odds ratio and corresponding 95% confidence interval. Odds ratios that are statistically significant (p-value < 0.05) are indicated with *, and those that are highly significant (p-value < 0.001) are indicated with ***.

## Notes

### Competing Interest Statement

PG, MN, CP, HB, UY, PL, PK, MN, JR, SA, and AV are employees of nference and have financial interests in the company and in the successful application of this research. VS is an employee of nference and Anumana and has financial interests in these companies and in the successful application of this research. JO has received small grants from nference, Inc., and personal consulting fees from Bates College and Elsevier Inc. All of these activities are outside of the present work. RH has received a small grant from nference, Inc. and Zealand Pharmaceuticals, and consulting fees from Nestle Nutrition. CR acknowledges funding from the National Institutes of Health (1OT2HL162096-01). AT is an employee and shareholder of Vir Biotechnology Inc.

### Funding Statement

This study was self-funded by nference.

### Author Declarations

This study was reviewed by the Mayo Clinic Institutional Review Board as a minimal-risk study and determined to be exempt. Participants were excluded if they did not have a research authorization on file. Further information on the Mayo Clinic Institutional Review Board and adherence to basic ethical principles underlying the conduct of research and ensuring that rights and well-being of potential research participants are adequately protected are available at the Mayo Clinic website.

